# Red flags to screen for vertebral or carotid artery dissection in patients with neck pain or headache: a scoping review protocol

**DOI:** 10.1101/2024.04.24.24306306

**Authors:** Daniel Feller, Annemarie de Zoete, Filippo Maselli, Firas Mourad, Alessandro Chiarotto, Bart Koes

## Abstract

**Background:** Vertebral and carotid artery dissections may present as neck pain and/or headache in their early phases. Consequently, clinicians must screen for arterial dissections when assessing neck pain and/or headache patients. Considering that no secondary studies have been published on the red flags to screen for arterial dissections in patients with neck pain/headache, our scoping review will aim to gain a comprehensive understanding of the existing literature on patients with a carotid or vertebral artery dissection presenting with a primary complaint of neck pain and/or headache regarding the prevalence of associated signs and symptoms (e.g., neurological impairments and visual problems), the pain characteristics (e.g., intensity and localization), the demographic characteristics (e.g., gender and age), the prevalence of risk factors (e.g., cardiovascular), the mechanism of onset, and other relevant clinical predictors (e.g., comorbidities).

**Methods:** We will search MEDLINE (via PubMed), Embase, CINHAL, and Scopus. In addition, we will use Web of Science to implement backward and forward citation tracking strategies. We will include any primary study design (e.g., case–control studies, case reports and case series, and cohort studies) written in English, Dutch, or Italian without any time restriction. To be included, studied had to focus on adult patients (> 18 years of age) of any gender with a diagnosis of vertebral or artery dissection and with a primary complaint of neck pain and/or headache, with a reporting on other signs/symptoms, pain characteristics, demographic information, risk factors, or onset mechanisms. Two authors will independently perform the study selection and data extraction phases. Results from the scoping review will be summarized descriptively through tables and diagrams. As a scoping review, we will highlight any gaps in the existing literature regarding our research questions.

## BACKGROUND

Although benign most of the time, neck pain and headache have been observed as the early manifestation of various serious pathologies. ^1,2^ One such pathology is vascular flow limitations of the cervico-cranial region, which can be defined as conditions that restrict or impair blood flow within the blood vessels supplying the head and neck area, namely the vertebrobasilar artery and the carotid arteries. ^3^ Depending on the severity and location of the vascular flow limitation, symptoms and signs can vary from dizziness, visual disturbances, weakness, and numbness, to stroke and hemorrhage. ^1^ Of all the pathologies causing vascular flow limitation in the cervico-cranial region, dissections (i.e., a tear in the inner layer of an artery) of the vertebral and carotid arteries are the ones that clinicians should be more aware of when assessing and treating patients presenting with a suspected musculoskeletal disorder of the cervico-cranial region. ^1,3^ In fact, the only early symptoms of dissections are usually neck pain and/or headaches, and, if left untreated, they could lead to cranial nerve palsies, Horner’s syndrome, or ischemic events like hindbrain strokes. ^4^ Therefore, it is essential for clinicians to screen for arterial dissections when assessing patients with neck pain and/or headaches by taking into consideration the presence of red flags, which are defined as signs, symptoms and patient characteristics that raise suspicion of serious spinal pathology. ^1,5^ Although the IFOMPT Cervical framework attempts to support clinicians in primary care ^1^, to date, there is no published secondary analysis on red flags for dissecting events of the carotid or vertebral arteries in patients presenting with a primary complaint of neck pain and/or headache.

## OBJECTIVES

Our objective is to gain a comprehensive understanding of the existing literature on patients with a carotid or vertebral artery dissection presenting with a primary complaint of neck pain and/or headache regarding the following aspects:

- The prevalence of associated signs and symptoms (e.g., neurological impairments and visual problems)
- The pain characteristics (e.g., intensity and localization)
- The demographic characteristics (e.g., gender and age)
- The prevalence of risk factors (e.g., cardiovascular)
- The mechanism of onset
- Other relevant clinical predictors (e.g., comorbidities)

## METHODS

The present scoping review will be conducted in accordance with the JBI methodology for scoping reviews. ^6^ The Preferred Reporting Items for Systematic Reviews and Meta-Analyses extension for Scoping Reviews (PRISMA-ScR) Checklist will be used for reporting the results. ^7^

### Inclusion criteria

Studies will be eligible for inclusion if they meet the following Population, Concept, and Context (PCC) criteria:

- Population. Adult patients (> 18 years of age) of any gender with a diagnosis of vertebral or artery dissection and with a primary complaint of neck pain and/or headache.
- Concept. Any primary study reporting data on other signs/symptoms, pain characteristics, demographic information, risk factors, or onset mechanisms.
- Context. Any clinical setting.

### Sources

This scoping review will consider any primary study design (e.g., case–control studies, case reports, and case series, cohort studies) written in English, Dutch, or Italian without any time restriction.

We will include studies with a mixed population (e.g., patients with dissections, but also other pathologies such as cervical cancer) if the majority of the population (> 80%) meets our inclusion criteria or the study presents a separate analysis for the patients who meet our inclusion criteria.

### Search strategy

We will search the following databases: MEDLINE (via Ovid), Embase, CINHAL (via EBSCO), and Web of Science. In addition, we will use Web of Science to implement backward and forward citation tracking strategies. All search strategies will be developed and implemented with the help of an expert librarian.

Following is reported the search string for MEDLINE (via Ovid):

*(exp Neck Pain / OR exp Headache/ OR exp Headache Disorders / OR Facial Pain / OR (((neck OR cervical* OR face OR facial*) ADJ3 pain*) OR cervicalgia* OR headache* OR head-ache* OR migraine*)*.*ab,ti,kw*.*) AND (Dissection, Blood Vessel / OR ((Carotid Artery Injuries / OR exp Carotid Arteries / OR Vertebral Artery /) AND (Dissection /)) OR Vertebral Artery Dissection / OR Carotid Artery, Internal, Dissection / OR ((carotid* OR vertebral* OR arter* OR cervic*) ADJ6 (dissect*))*.*ab,ti,kw. OR ((carotid* OR vertebral* OR arter* OR cervic*) AND (dissect*))*.*ti*.*) NOT (Coronary Artery Dissection, Spontaneous* .*nm. OR * Aortic Dissection / OR * Acute Coronary Syndrome / OR (((coronary OR aort*) ADJ3 dissect*) OR (acute ADJ3 coronar* ADJ3 syndrome*))*.*ti*.*) NOT (exp animals/ NOT humans/) AND (english*.*la. OR dutch*.*la. OR Italian*.*la*.*) NOT (Systematic Review / OR Meta-Analysis / OR ((systematic* ADJ3 review*) OR (meta-analys*))*.*ti*.)

### Study selection

Duplicates will be removed using EndNote. Two researchers will independently perform the study selection process, firstly by title/abstract and secondly by full text. Any disagreement will be resolved by consensus or by the decision of a third author. We will use the online electronic systematic review software package (Rayyan QCRI) to organize and track the selection process. ^4^ Reasons for the exclusion of any full-text source of evidence will be recorded and reported.

### Data extraction

The data extraction process will be conducted independently by two reviewers using a standardized form. Any discrepancies will be resolved by a consensus between the two authors and eventually by a third author’s decision. We will aim to extract the following information:

- First author and publication year
- Study design
- Clinical setting (patient population source, country)
- Characteristics of the neck pain and/or headache (e.g., duration of neck pain, pain location)
- Other reported signs and symptoms (e.g., neurological signs)
- Demographic characteristics of the included patients (e.g., age
- Prevalence of cardiovascular risk factors
- Mechanisms of onset
- Other relevant clinical predictors (e.g., comorbidities)
- Diagnostic procedure used to make the diagnosis of arterial dissection (e.g., imaging, IFOMPT framework)

The process of charting in scoping reviews is iterative. If additional relevant items emerge during full-text analysis, we will extract more data. Any modifications to the data extraction phase will be mentioned in the full manuscript. Relevant missing data will be gathered by contacting the corresponding author with a maximum of two attempts on a weekly basis.

### Data synthesis

Results from the scoping review will be summarized descriptively through tables and diagrams. Specifically, we will narratively summarize and synthesize data relating to the prevalence of associated signs and symptoms, pain characteristics, demographic information, the prevalence of cardiovascular risk factors, and the mechanisms of onset. As a scoping review, we will highlight any gaps in the existing literature regarding our research questions.

## Data Availability

All data produced in the present study will be available upon reasonable request to the authors.

